# Waning Effectiveness of the BNT162b2 Vaccine Against Infection in Adolescents

**DOI:** 10.1101/2022.01.04.22268776

**Authors:** Ottavia Prunas, Daniel M. Weinberger, Virginia E. Pitzer, Sivan Gazit, Tal Patalon

## Abstract

**Background:** The short-term effectiveness of a two-dose regimen of the BioNTech/Pfizer mRNA BNT162b2 vaccine for adolescents has been demonstrated. However, little is known about the long-term effectiveness in this age group. It is known, though, that waning of vaccine-induced immunity against infection in adult populations is evident within a few months.

**Methods:** Leveraging the centralized computerized database of Maccabi Healthcare Services (MHS), we conducted a matched case-control design for evaluating the association between time since vaccination and the incidence of infections, where two outcomes were evaluated separately: a documented SARS-CoV-2 infection (regardless of symptoms) and a symptomatic infection (COVID-19). Cases were defined as individuals aged 12 to 16 with a positive PCR test occurring between June 15 and December 8, 2021, when the Delta variant was dominant in Israel. Controls were adolescents who had not tested positive previously.

**Results:** We estimated a peak vaccine effectiveness between 2 weeks and 3 months following receipt of the second dose, with 85% and 90% effectiveness against SARS-CoV-2 infection and COVID-19, respectively. However, in line with previous findings for adults, waning of vaccine effectiveness was evident in adolescents as well. Long-term protection conferred by the vaccine was reduced to 75-78% against infection and symptomatic infection, respectively, 3 to 5 months after the second dose, and waned to 58% against infection and 65% against COVID-19 after 5 months.

**Conclusions:** Like adults, vaccine-induced protection against both SARS-CoV-2 infection and COVID-19 wanes with time, starting three months after inoculation and continuing for more than five months.

## Introduction

The BioNTech/Pfizer mRNA BNT162b2 vaccine was approved for adolescents by the United States Food and Drug Administration (FDA) and the European Medicines Agency in May 2021.^1,2^ Shortly after, in June 2021, Israel, an early adopter of the vaccination campaign in adults, launched a vaccination campaign for adolescents,^3^ and has since continued to promote it.^4^

The short-term effectiveness of a two-dose regimen of the BioNTech/Pfizer mRNA BNT162b2 vaccine for adolescents has been demonstrated both in clinical trials^5^ and real-world studies.^6^ However, little is known about the long-term effectiveness of the vaccine in this age group. It is known, though, that waning of vaccine-induced immunity of the BNT162b2 vaccine in adult populations is evident within a few months of administration,^7–11^ though protection against severe disease is more sustained.^12^ The question of waning immunity in the adolescent population has important implications for future areas of vaccine research and policy. For example, the US FDA and Israel have already extended the eligibility of the booster (third) dose for ages 12-15,^13,14^ and other countries are considering this decision.

To address this issue, we conducted a retrospective matched case-control study aimed at evaluating the duration of protection conferred by the BNT162b2 vaccine on adolescents aged 12 to 16, leveraging data from Maccabi Healthcare Services (MHS), Israel’s second largest Health Maintenance Organization, which covers 2.5 million members. The 6-month follow-up period of the study, from June 15 to December 8, 2021, represents the longest published on this age group to date, and corresponds to a time when the Delta (B.1.617.2) variant was dominant in Israel, prior to the surge of the Omicron variant.^15^

## Methods

### Data sources

MHS is a 2.5-million-member, not-for-profit health-fund in Israel. It is the second largest in Israel, covering 26.7% of the population and providing a representative sample of the Israeli population. MHS has maintained a centralized database of Electronic Medical Records (EMRs) for three decades, with less than 1% disengagement rate among its members, allowing for a comprehensive longitudinal medical follow-up. The centralized dataset includes extensive demographic data, clinical measurements and evaluations, outpatient and hospital diagnoses and procedures, medications dispensed, imaging performed and comprehensive laboratory data from a single central laboratory.

### Study population and data collection

The study population consisted of MHS members, aged 12-16 years, who received either one or two doses of the BNT162b2 vaccine. Anonymized EMRs were retrieved from MHS’ centralized computerized database. Analyses focused on the period from June 15, 2021, to December 8, 2021, when the Delta variant was dominant in Israel. Participants were excluded from the study if they tested positive for SARS-CoV-2 by a polymerase chain reaction (PCR) test before the start of the follow-up period or disengaged from MHS for any reason prior to the study period.

Individual-level data for the study population included age, biological sex, and a coded geographical statistical area (GSA; the smallest geostatistical unit of the Israeli census, which correspond to neighborhoods assigned by Israel’s National Bureau of Statistics). Data collected also encompassed the last documented body mass index (BMI) (where obesity was defined as BMI ≥ 30). COVID-19-related information included dates of the first and second dose of the BNT162b2 vaccine and results of any PCR tests for SARS-CoV-2, including tests performed outside of MHS. Additionally, the dataset contained information on symptoms for some of the participants who tested positive. The information about COVID-19-related symptoms was extracted from EMRs, where they were recorded by the primary care physician or a certified nurse who conducted in-person or phone visits with each infected individual. Information on symptoms was only available for individuals who had a positive test for SARS-CoV-2. For individuals who had multiple positive tests, the date of diagnosis was defined as the date of the first positive PCR.

#### Statistical analysis

Two main outcomes were evaluated: SARS-CoV-2 infection (regardless of the presence of symptoms) and symptomatic SARS-CoV-2 infection (COVID-19).

##### Analysis 1: matched case-control analysis for breakthrough infections

We used a matched case-control design^16–18^ for evaluating the association between time since vaccination and the incidence of infections. Cases were defined as individuals with a positive PCR test occurring after June 15, 2021, among those 12-16 years of age who did not have a previous positive test recorded. Eligible controls were individuals who had not tested positive prior to the date of the positive PCR of their matched case. Controls were matched by residential socioeconomic status and biological sex. Up to 20 controls per case were drawn from the entire population; 90% of the cases were matched to 20 controls, 96% of the cases had at least 10 controls, and 99% of the cases had at least 5 controls).

The analysis sought to estimate the reduction in the odds of a positive test at different time intervals following receipt of the first and second vaccine doses (0-6 days following the first dose; 7 days after dose 1 to 13 days after dose 2; and 14-89 days, 90-149 days and 150-180 days following the second dose). The 7-day and 14-day cutoffs for the first and second doses were chosen based on previous research on vaccine effectiveness in adults.^19,20^ The reference group in the analysis was those who were unvaccinated. We included obesity (i.e., BMI ≥ 30) as a covariate, which has been linked to severity of COVID-19 symptoms.^21–23^ We also tested our model by including an additional covariate consisting of the number of tests performed before the study period, i.e. between March 1, 2020, to June 14, 2021, to adjust for potential differences in health-seeking behavior.^16^ The rationale is that there is a correlation between the number of tests before the study period and the one during it, potentially correcting for a possible detection bias.^7,9^ However, repeated testing was significantly lower compared to adults,^7,16,24^ and results were not materially changed.

A conditional logistic regression model was fit to the data. The vaccine effectiveness (VE) of the first and second doses (compared to being unvaccinated) was calculated as 100%*[1-(Odds Ratio)] for each of the time-since-vaccination categories.

Additionally, we carried out a complementary analysis, utilizing a test-negative design. Details can be found in the Supplementary Appendix.

##### Analysis 2: matched case-control analysis for symptomatic breakthrough infections

Similar to the analysis of infections, we performed a matched case-control analysis for symptomatic infection (COVID-19). Cases were individuals who tested positive and exhibited COVID-19-related symptoms after June 15, 2021. Eligible controls were individuals who had not tested positive. Matching was performed as for the infection case-control analysis. Again, up to 20 controls per case were drawn from the entire population; 89% of cases were matched to 20 controls, 96% of cases had at least 10 controls, and 99% of cases had at least 5 controls). A conditional logistic regression was fit to the data, adjusting for obesity, as specified in analysis 1.

Analyses were performed using R version 4.0.5. The analysis conformed to the STROBE checklist for case-control studies.

#### Ethics declaration

This study was approved by the MHS (Maccabi Healthcare Services) Institutional Review Board. Due to the retrospective design of the study, informed consent was waived by the IRB, and all identifying details of the participants were removed before computational analysis.

## Results

During the follow-up period, 274,431 PCR tests were performed among 129,909 MHS members 12-16 years of age who did not have a previous documented infection. Baseline characteristics of the participants are given in Table 1. Overall, vaccinated individuals had a notably lower percent of tests that were positive; 6.6% of tests among unvaccinated individuals were positive, compared with <1.4% for those who received their second dose 14-149 days before the test, and 3.6% for those who received their second dose ≥150 days before the test (Table 2). There were 14 hospitalizations for COVID-19 and no deaths reported.

**Table 1.** Demographic characteristics of individuals who were tested between June 15 and December 8, 2021

| **Characteristics** | **Control**<br>(n=226,201) | **Case**<br>(n=11,822) | **Overall**<br>(n=238,023) |
| --- | --- | --- | --- |
| **Age years, mean (SD)** | 14.0 (1.4) | 13.7 (1.4) | 14.0 (1.4) |
| **Sex – no. (%)** |  |  |  |
| Female | 117,697 (52.0%) | 6143 (52.0%) | 123,840<br>(52.0%) |
| Male | 108,504 (48.0%) | 5679 (48.0%) | 114,183 (48.0%) |
| **Comorbidities – no. (%)** |  |  |  |
| Obesity (BMI $\geq 30$) | 7633 (3.4%) | 327 (2.8%) | 7,960 (3.3%) |
| **Socioeconomic status by GSA** |  |  |  |
| High (7-10) | 94,810 (41.9%) | 4902 (41.5%) | 99,712 (41.9%) |
| Medium (5-6) | 82,813 (36.6%) | 4360 (36.9%) | 87,173 (36.6%) |
| Low (0-4) | 45,034 (19.9%) | 2377 (20.1%) | 47,411 (19.9%) |
| Missing | 3544 (1.6%) | 183 (1.5%) | 3727 (1.6%) |

SD – Standard Deviation; BMI – Body Mass Index; GSA – Geographical Statistical Area.

**Table 2.**
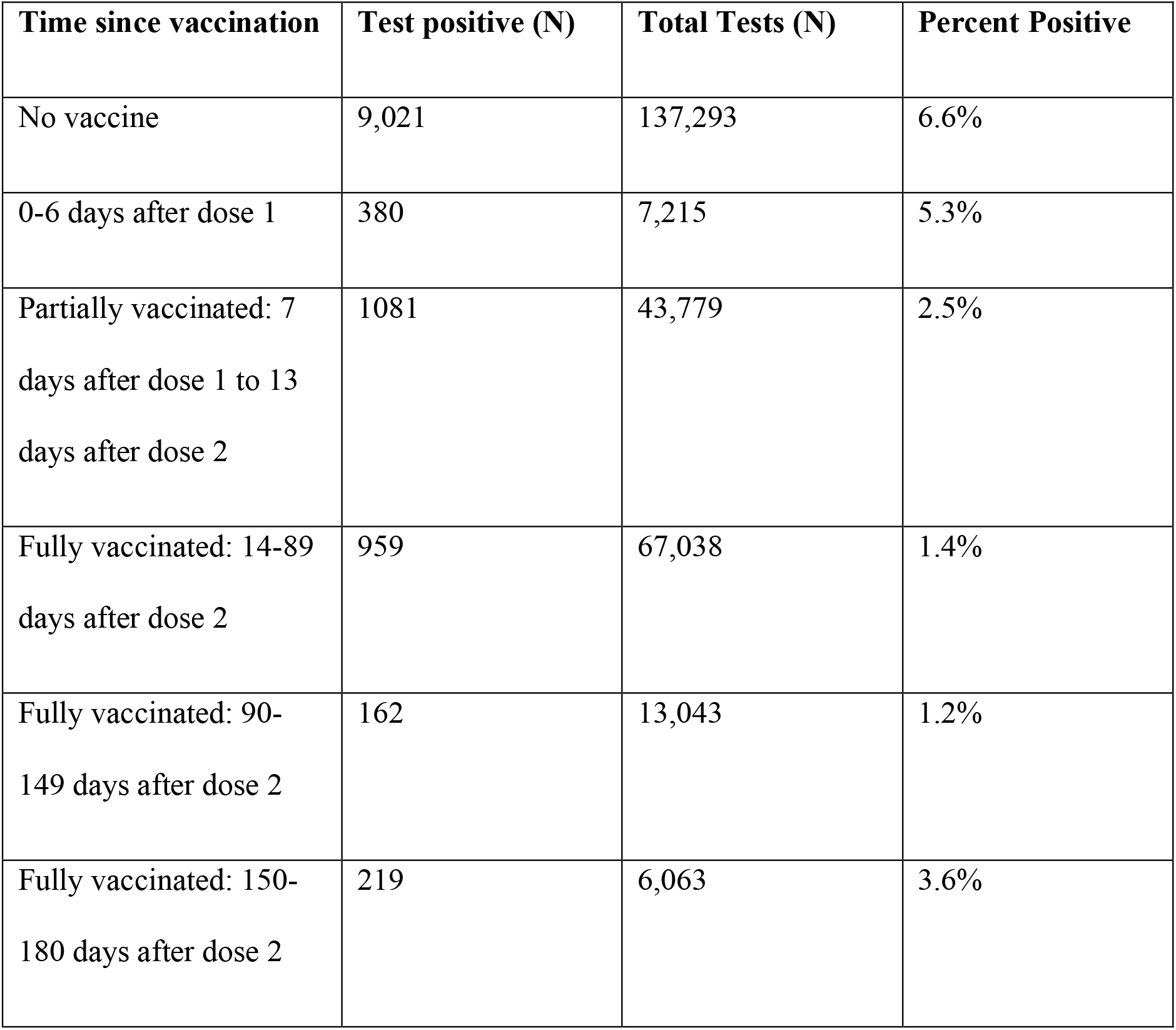
Number of SARS-CoV-2-positive tests and total number of tests among adolescents by vaccination category for the study period, June 15 - December 8, 2021.

| **Time since vaccination** | **Test positive (N)** | **Total Tests (N)** | **Percent Positive** |
| --- | --- | --- | --- |
| No vaccine | 9,021 | 137,293 | 6.6% |
| 0-6 days after dose 1 | 380 | 7,215 | 5.3% |
| Partially vaccinated: 7 days after dose 1 to 13 days after dose 2 | 1081 | 43,779 | 2.5% |
| Fully vaccinated: 14-89 days after dose 2 | 959 | 67,038 | 1.4% |
| Fully vaccinated: 90-149 days after dose 2 | 162 | 13,043 | 1.2% |
| Fully vaccinated: 150-180 days after dose 2 | 219 | 6,063 | 3.6% |

### Vaccine effectiveness against breakthrough infections

The effectiveness of the first and second doses compared to the unvaccinated population initially increased over time following receipt of the vaccine, with a small reduction in the odds of testing positive in days 0-6 following the first dose (VE=12%, 95% CI: 2%, 21%), moderate effectiveness for the period from day 7 after the first dose to day 13 after the second dose (VE=52%, 95% CI: 49%, 55%), and high effectiveness in the period from days 14-89 following dose 2 (VE=85%, 95% CI: 84%, 86%) (Table S3, Figure 1). The effectiveness of the vaccine against infection was subsequently reduced to 75% (95% CI: 71%, 79%) and 58% (95% CI: 52%, 64%) after 90-149 days and 150-180 days following receipt of the second dose, respectively. The results of the test-negative design showed a similar decline in vaccine effectiveness (Table S3).

**Figure 1.**
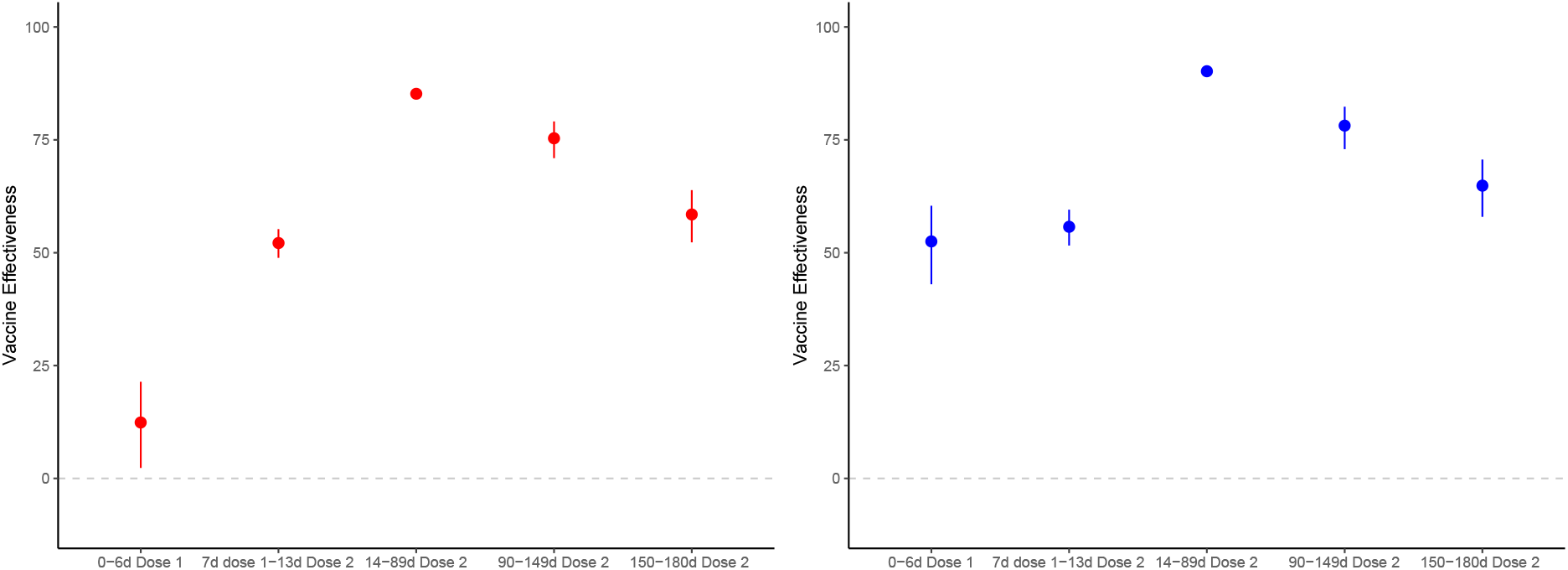
Reduction in the odds of testing positive for SARS-CoV-2 (left panel) and in having a positive test with symptoms (right panel) among individuals who had received one or two doses of BNT162b2 vaccine compared to unvaccinated adolescents, by time since vaccination. In red, vaccine effectiveness against SARS-CoV-2 infection, and in blue, against symptomatic infection. Vertical bars represent 95% confidence intervals.

### Vaccine effectiveness against symptomatic breakthrough infections

We next evaluated vaccine effectiveness against symptomatic infection, comparing vaccinated to unvaccinated individuals. In those partially vaccinated (i.e., persons between 7 days after the first dose up to 13 days after the second dose), we observed a 56% (95% CI: 52%, 60%) reduction in the odds of having a symptomatic SARS-CoV-2 infection compared to unvaccinated adolescents. This reduction was more marked 14-89 days following the second dose, where the vaccine effectiveness against COVID-19 was 90% (95% CI: 89%, 91%) (Table S2). Similar to the analysis of all infections (regardless of symptomatic presentation), the estimated effectiveness of the vaccine against COVID-19 subsequently decreased, with VE=78% (95% CI: 73%, 82%) for days 90-149 and VE=65% (95% CI: 58%, 71%) for days 150-180 after the second dose.

## Discussion

In this study, we found that the BioNTech/Pfizer mRNA BNT162b2 vaccine provided strong short-term protection against any SARS-CoV-2 infection and symptomatic infection (COVID-19) with the Delta variant in adolescents, confirming previous findings.^5,6^ Using a matched case-control analysis, we estimated a peak vaccine effectiveness between 2 weeks and 3 months following receipt of the second dose, with 85% and 90% effectiveness against SARS-CoV-2 infection and COVID-19, respectively. However, in line with previous findings for adults,^7–9^ waning of vaccine effectiveness was evident in adolescents as well. Long-term protection conferred by the vaccine was reduced to 75-78% against infection and symptomatic infection, respectively, 3 to 5 months after the second dose, and waned to 58% against infection and 65% against symptomatic infection after 5 months.

We evaluated vaccine effectiveness against infection and COVID-19, but did not evaluate effectiveness against severe disease. Even in the absence of vaccination, adolescents have a much lower risk of hospitalization and death compared to adults,^25,26^ and the number of events in this population was small, though the nature and risk of persistent symptoms (i.e. long COVID) is still being characterized.^27^ Other studies in adults have found that vaccine effectiveness against severe outcomes has been maintained at higher levels than effectiveness against infection.^12^ Decisions to vaccinate and to use a booster dose among adolescents, as well as prioritization of vaccines among different age and risk groups, will therefore depend on the policy goals, i.e. reducing transmission of SARS-CoV-2 in the population in the short term versus reducing the burden of disease in the population.

This study has several limitations. The estimates of vaccine-induced protection against symptomatic infection should be interpreted with caution, since a protective effect of the vaccine was already evident a few days after the receipt of the first dose (52% effectiveness at 0-6 days). We did not expect to find a significant effect of vaccination in the first week after receipt of the first dose, since it presumably takes time for the vaccine to induce an immune response. We observed a small but significant reduction in the odds of infection in the six days following the first dose and a larger reduction in the odds of symptomatic infection, which could indicate a potential bias. It is possible that individuals are less likely to be tested immediately after vaccination (e.g. because they attribute symptoms to temporary side effects), and are therefore less likely to be detected (rather than less likely to be infected) compared to unvaccinated individuals, especially symptomatic ones. However, this bias would most likely be short-lived, so the estimates of vaccine effectiveness against COVID-19 could be more reliable a couple of weeks after receipt of the first dose.^9,16^

A second limitation is with regards the generalizability of our findings in light of the emergence of novel SARS-CoV-2 variants. Our study only estimates the short- and long-term vaccine effectiveness against the Delta variant, which was the dominant strain in Israel at the time of our study. The protection conferred against other strains, including the Omicron variant, cannot be inferred.

### Conclusion

Our analyses suggest that, compared to those who are unvaccinated, adolescents aged 12 to 16 who received two doses of the BNT162b2 vaccine have a lower risk of contracting SARS-CoV-2 infection, as detected by PCR, and a lower risk of symptomatic infection. However, like adults, vaccine-induced protection against both SARS-CoV-2 infection and symptomatic infection wanes with time, starting three months after inoculation and continuing for more than five months.

## Data Availability

According to the Israel Ministry of Health regulations, individual-level data cannot be shared openly. Specific requests for remote access to de-identified community-level data should be referred to KSM, Maccabi Healthcare Services Research and Innovation Center.

## Code availability statement

Specific requests for remote access to the code used for data analysis should be referred to KSM, Maccabi Healthcare Services Research and Innovation Center.

## Supplementary Appendix

### Supplementary Analysis: Test-Negative Design

As a secondary analysis, we ran a test-negative design, where ‘cases’ were defined as those who had a positive PCR test for SARS-CoV-2 and ‘controls’ were those testing negative. Individuals could contribute multiple negative tests to the analysis, but were excluded once they tested positive. If multiple tests were conducted within a 5-day period, the day of test was considered to be the first day of that test series. Similar to the matched case-control analysis, we estimated the reduction in the odds of a positive test at different time intervals following receipt of the first and second vaccine doses (0-6 days following the first dose; 7 days after dose 1 to 13 days after dose 2; and 14-89 days, 90-149 days and 150-180 days following the second dose). We also included obesity (i.e., Body Mass Index ≥ 30) as a covariate. The number of positive tests performed on that day throughout the entire population (log-transformed) was included as an additional covariate to adjust for potential variability in levels of exposure at different time points.

A generalized estimating equation (GEE) logistic regression model was fit to the data. The benefit of the first and second dose (compared to those unvaccinated) was calculated as 100%*[1-(Odds Ratio)] for each of the time-since-vaccination categories. This is analogous to a standard vaccine effectiveness estimate, but adjusting for vaccination history. The GEE model accounts for repeated samples collected from the same individual over time and assumed an exchangeable correlation structure. Results comparing the test-negative design with the matched case-control analysis, i.e. our primary analysis, are presented in Table S3.

**Table S1.**
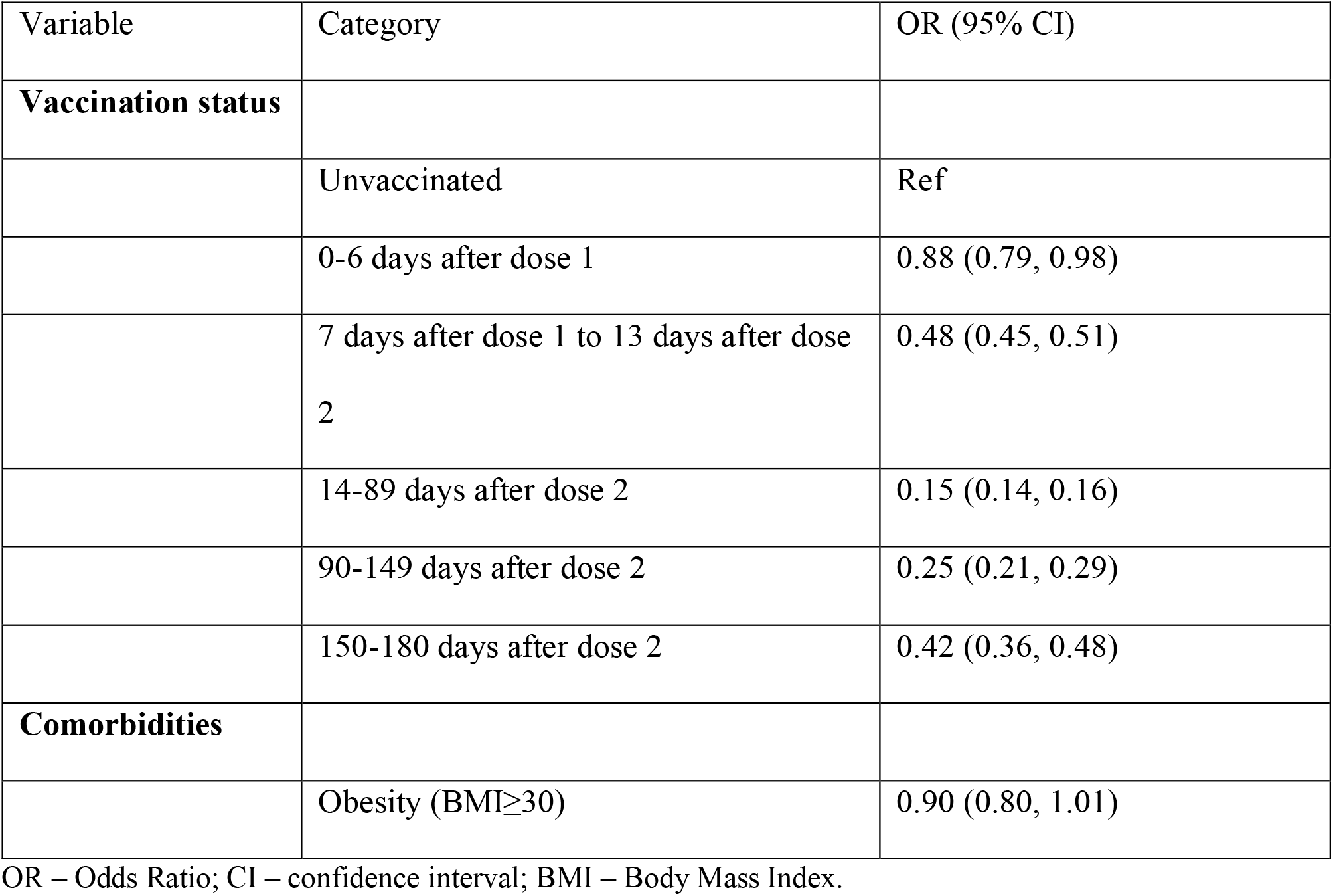
Odds ratios for SARS-CoV-2 infection, based on vaccination category, demographics and comorbidities, for the matched case-control design.

| Variable | Category | OR (95% CI) |
| --- | --- | --- |
| **Vaccination status** |  |  |
|  | Unvaccinated | Ref |
|  | 0-6 days after dose 1 | 0.88 (0.79, 0.98) |
|  | 7 days after dose 1 to 13 days after dose 2 | 0.48 (0.45, 0.51) |
|  | 14-89 days after dose 2 | 0.15 (0.14, 0.16) |
|  | 90-149 days after dose 2 | 0.25 (0.21, 0.29) |
|  | 150-180 days after dose 2 | 0.42 (0.36, 0.48) |
| **Comorbidities** |  |  |
| | Obesity (BMI≥30) | 0.90 (0.80, 1.01) |

OR – Odds Ratio; CI – confidence interval; BMI – Body Mass Index.

**Table S2.** Odds ratios for symptomatic SARS-CoV-2 infection, based on vaccination category, demographics and comorbidities, for the matched case-control design.

| Variable | Category | OR (95% CI) |
| --- | --- | --- |
| **Vaccination status** |  |  |
|  | Unvaccinated | Ref |
|  | 0-6 days after dose 1 | 0.48 (0.40, 0.57) |
|  | 7 days after dose 1 to 13 days after dose 2 | 0.44 (0.40, 0.48) |
|  | 14-89 days after dose 2 | 0.10 (0.09, 0.11) |
|  | 90-149 days after dose 2 | 0.22 (0.18, 0.27) |
|  | 150-180 days after dose 2 | 0.35 (0.29, 0.42) |
| **Comorbidities** |  |  |
| | Obesity (BMI≥30) | 0.98 (0.85, 1.13) |

OR – Odds Ratio; CI – confidence interval; BMI – Body Mass Index.

**Table S3.** Vaccine effectiveness of one and two doses of BNT162b2 against SARS-CoV-2 infection using both a matched case-control and a test-negative design.

| **Time since vaccination** | **Matched case-control design VE (95% CI)** | **Test-negative design VE (95% CI)** |
| --- | --- | --- |
| 0-6 days after dose 1 | 12% (2%, 21%) | 19% (10%, 27%) |
| 7 days after dose 1 to<br>13 days after dose 2 | 52% (49%, 55%) | 62% (60%, 64%) |
| 14-89 days after dose 2 | 85% (84%, 86%) | 84% (82%, 85%) |
| 90-149 days after dose 2 | 75% (71%, 79%) | 67% (61%, 72%) |
| 150-180 days after dose 2 | 58% (52%, 64%) | 50% (43%, 57%) |

VE – vaccine effectiveness; CI – confidence interval

## References

1. Food and Drug Administration (FDA). Coronavirus (COVID-19) Update: FDA Authorizes Pfizer-BioNTech COVID-19 Vaccine for Emergency Use in Adolescents in Another Important Action in Fight Against Pandemic [Internet]. Food Drug Adm. 2021 [cited 2021 May 18];Available from: https://www.fda.gov/news-events/press-announcements/coronavirus-covid-19-update-fda-authorizes-pfizer-biontech-covid-19-vaccine-emergency-use

2. First COVID-19 vaccine approved for children aged 12 to 15 in EU | European Medicines Agency [Internet]. [cited 2021 Dec 31];Available from: https://www.ema.europa.eu/en/news/first-covid-19-vaccine-approved-children-aged-12-15-eu

3. Ministry of Health’s Position Regarding the Expansion of the Vaccination Operation to Ages 12-16 Years Ministry of Health [Internet]. [cited 2021 Dec 31];Available from: https://www.gov.il/en/departments/news/02062021-01

4. Campaign to Motivate Teenagers to Get Vaccinated | Ministry of Health [Internet]. [cited 2022 Jan 1];Available from: https://www.gov.il/en/Departments/news/24082021-04

5. Frenck RW, Klein NP, Kitchin N, et al. Safety, Immunogenicity, and Efficacy of the BNT162b2 Covid-19 Vaccine in Adolescents. N Engl J Med [Internet] 2021 [cited 2021 Dec 31];385(3):239–50. Available from: https://www.nejm.org/doi/full/10.1056/NEJMoa2107456

6. Glatman-Freedman A, Hershkovitz Y, Kaufman Z, Dichtiar R, Keinan-Boker L, Bromberg M. Effectiveness of BNT162b2 Vaccine in Adolescents during Outbreak of SARS-CoV-2 Delta Variant Infection, Israel, 2021. Emerg Infect Dis [Internet] 2021 [cited 2021 Dec 31];27(11):2919. Available from: /pmc/articles/PMC8544958/

7. Mizrahi B, Lotan R, Kalkstein N, et al. Correlation of SARS-CoV-2-breakthrough infections to time-from-vaccine. Nat Commun 2021 121 [Internet] 2021 [cited 2021 Nov 14];12(1):1–5. Available from: https://www.nature.com/articles/s41467-021-26672-3

8. Chemaitelly H, Tang P, Hasan MR, et al. Waning of BNT162b2 Vaccine Protection against SARS-CoV-2 Infection in Qatar. N Engl J Med [Internet] 2021 [cited 2021 Dec 28];385(24):e83. Available from: https://www.nejm.org/doi/full/10.1056/NEJMoa2114114

9. Bar-On YM, Goldberg Y, Mandel M, et al. Protection of BNT162b2 Vaccine Booster against Covid-19 in Israel. N Engl J Med [Internet] 2021;385(15):1393–400. Available from: https://doi.org/10.1056/NEJMoa2114255

10. Levin EG, Lustig Y, Cohen C, et al. Waning Immune Humoral Response to BNT162b2 Covid-19 Vaccine over 6 Months. N Engl J Med [Internet] 2021 [cited 2021 Dec 28];385(24):e84. Available from: https://www.nejm.org/doi/full/10.1056/NEJMoa2114583

11. Levine-Tiefenbrun M, Yelin I, Alapi H, et al. Viral loads of Delta-variant SARS-CoV-2 breakthrough infections after vaccination and booster with BNT162b2. Nat Med 2021 [Internet] 2021 [cited 2021 Nov 14];1–3. Available from: https://www.nature.com/articles/s41591-021-01575-4

12. Tenforde MW, Self WH, Naioti EA, et al. Sustained Effectiveness of Pfizer-BioNTech and Moderna Vaccines Against COVID-19 Associated Hospitalizations Among Adults — United States, March–July 2021. Morb Mortal Wkly Rep [Internet] 2021 [cited 2021 Dec 28];70(34):1156. Available from: /pmc/articles/PMC8389395/

13. F.D.A. Plans to Allow 12-to 15-Year-Olds to Receive Pfizer Boosters -The New York Times [Internet]. [cited 2022 Jan 1];Available from: https://www.nytimes.com/2021/12/30/us/politics/boosters-12-15-year-olds-omicron.html

14. The Vaccination Committee Recommends: 3 Weeks Between the First and Second Dose Also for Children 5-11 | Ministry of Health [Internet]. [cited 2022 Jan 1];Available from: https://www.gov.il/en/departments/news/21112021-04

15. SARS-CoV-2 variants in analyzed sequences, Israel [Internet]. [cited 2021 Dec 30];Available from: https://ourworldindata.org/grapher/covid-variants-area?country=~ISR

16. Patalon T, Gazit S, Pitzer VE, et al. Odds of Testing Positive for SARS-CoV-2 Following Receipt of 3 vs 2 Doses of the BNT162b2 mRNA Vaccine. JAMA Intern Med [Internet] 2021 [cited 2021 Dec 3];Available from: https://jamanetwork.com/journals/jamainternalmedicine/fullarticle/2786890

17. V E, RW P, M A, L P. Comparison of nested case-control and survival analysis methodologies for analysis of time-dependent exposure. BMC Med Res Methodol [Internet] 2005 [cited 2021 Aug 28];5(1). Available from: https://pubmed.ncbi.nlm.nih.gov/15670334/

18. Zakeri R, Bendayan R, Ashworth M, et al. A case-control and cohort study to determine the relationship between ethnic background and severe COVID-19. EClinicalMedicine [Internet] 2020 [cited 2021 Aug 28];28. Available from: http://www.thelancet.com/article/S2589537020303187/fulltext

19. Chodick G, Tene L, Rotem RS, et al. The Effectiveness of the Two-Dose BNT162b2 Vaccine: Analysis of Real-World Data. Clin Infect Dis 2021;

20. Dagan N, Barda N, Kepten E, et al. BNT162b2 mRNA Covid-19 Vaccine in a Nationwide Mass Vaccination Setting. N Engl J Med [Internet] 2021 [cited 2021 Apr 20];384(15). Available from: https://pubmed.ncbi.nlm.nih.gov/33626250/

21. Gao F, Zheng KI, Wang XB, et al. Obesity Is a Risk Factor for Greater COVID-19 Severity. Diabetes Care [Internet] 2020 [cited 2021 Dec 31];43(7):e72–4. Available from: https://doi.org/10.2337/dc20-0682

22. Simonnet A, Chetboun M, Poissy J, et al. High prevalence of obesity in severe acute respiratory syndrome coronavirus-2 (SARS-CoV-2) requiring invasive mechanical ventilation. Obesity (Silver Spring) [Internet] 2020 [cited 2021 Dec 31];28(7):1195–9. Available from: /pmc/articles/PMC7262326/?report=abstract

23. Alberca RW, Oliveira L de M, Branco ACCC, Pereira NZ, Sato MN. Obesity as a risk factor for COVID-19: an overview. https://doi.org/101080/1040839820201775546 [Internet] 2020 [cited 2021 Dec 31];61(13):2262–76. Available from: https://www.tandfonline.com/doi/abs/10.1080/10408398.2020.1775546

24. Goldberg Y, Mandel M, Bar-On YM, et al. Waning Immunity after the BNT162b2 Vaccine in Israel. N Engl J Med [Internet] 2021 [cited 2022 Jan 2];385(24):e85. Available from: https://www.nejm.org/doi/full/10.1056/NEJMoa2114228

25. Smith C, Odd D, Harwood R, et al. Deaths in children and young people in England after SARS-CoV-2 infection during the first pandemic year. Nat Med [Internet] 2021 [cited 2022 Jan 4];1–8. Available from: https://www.nature.com/articles/s41591-021-01578-1

26. Saxena S, Skirrow H, Wighton K. Should the UK vaccinate children and adolescents against covid-19? BMJ [Internet] 2021 [cited 2022 Jan 4];374. Available from: https://www.bmj.com/content/374/bmj.n1866

27. Zimmermann P, Pittet LF, Curtis N. How Common is Long COVID in Children and Adolescents? Pediatr Infect Dis J [Internet] 2021 [cited 2022 Jan 4];40(12):e482. Available from: /pmc/articles/PMC8575095/

